# Factors associated with underutilization of cervical cancer screening services among HIV positive women in Serenje District, Central Province

**DOI:** 10.1101/2025.08.05.25333091

**Authors:** Maphious Shindanyi Makina, Phoebe Albina Bwembya, Cosmas Zyambo, Alice Ngoma Hazemba

**Author notes:** Corresponding author: Correspondence to Maphious Shindanyi Makina.

## Abstract

Cervical cancer is the most common cancer among women living with Human Immunodeficiency Virus (HIV) (UNAIDS, 2019^59^). An estimated 604,000 new cases were diagnosed globally, with 342,000 deaths recorded in 2020 (Sung et al., 2021^69^). According to WHO (2020), approximately 6.5% of all female cancers are cervical cancer-related, with 5% of cases attributable to HIV. Zambia ranks second in cervical cancer incidence, with an estimated mortality of 1,839 (WHO, 2020^56^). The current utilization of cervical cancer screening services (CCSS) in Zambia among HIV-positive women stands at 27%, indicating underutilization (UNAIDS, 2019^59^). In Serenje District, only 25.8% of HIV-positive women access these services. This study determined factors associated with the underutilization of CCSS among HIV-positive women in Serenje District.

**Methods:** A descriptive cross-sectional study was conducted involving 303 HIV-positive women in Serenje District of Central Province, Zambia. Systematic random sampling was used to recruit respondents from selected facilities, and a structured questionnaire was employed for data collection. Statistical Software (STATA V15) was utilized for analysis. Univariate, bivariate, and multivariable logistic regressions were performed to determine associations between variables.

**Results:** Utilization of CCSS in Serenje District was low, with only 42.24% of respondents reporting service use. Being far from a healthcare facility [AOR: 0.49 (P<0.036, 95% CI 0.25-0.96)], low/inadequate knowledge [AOR: 0.26 (P<0.001, 95% CI 0.13-0.54)], low income [AOR: 0.16 (P<0.001, 95% CI 0.06-0.37)], and negative attitudes towards CCSS [AOR: 0.26 (P<0.001, 95% CI 0.12-0.53)] were significant contributors to reduced utilization.

**Conclusion:** Low knowledge, poor attitudes, far distances to healthcare facilities, and poor socioeconomic status were identified as key factors contributing to the underutilization of CCSS. It is recommended that the district designs and implements awareness campaigns on cervical cancer screening services, engages in outreach awareness campaigns, and conducts screening camps to bring these services closer to the communities.

## BACKGROUND

Cervical cancer is the most common cancer among women living with HIV. It is preventable and curable if diagnosed and treated early (UNAIDS, 2019^59^). Approximately 70% of all cervical cancers and pre-cancerous cervical lesions are caused by sexually transmitted Human Papillomavirus (HPV) types 16 and 18 (WHO, 2020^72^).

According to WHO (2020^56^), an estimated 604,000 women were diagnosed with cervical cancer globally. About 6.5% of all female cancers and 5% of cases are attributable to HIV. HIV accounts for more than 40% of cervical cancer cases in eight sub-Saharan African countries. Despite their increased risk, women living with HIV in sub-Saharan Africa continue to experience limited access to regular screening for cervical cancer (UNAIDS, 2019^59^).

In 2020, 85% of sub-Saharan African women with cervical cancer were reported to have HIV (WHO, 2020^27^). Countries with the highest rates of cervical cancer by age-standardized incidence per 100,000 include Swaziland (75.3%), Malawi (72.9%), and Zambia (66.4%) (Bray, 2018^21^). The WHO recommends simple, low-cost visual inspection with acetic acid (VIA) as the optimal screening method for developing countries. However, utilization of screening services among women living with HIV aged 30-49 years remains low in Malawi (19%) and Zambia (27%) (UNAIDS, 2019^59^).

In Zambia, cervical cancer represents 34.3% of all cancers, making it the most prevalent cancer in the country (Kalubula et al., 2018^20^). The incidence rate of cervical cancer is 58.4 per 100,000, with a mortality rate of 36.2 per 100,000 (Venturas et al., 2017^19^), which remain unacceptably high. The Ministry of Health in Zambia launched cervical cancer screening services in 51 hospitals nationwide to reduce cervical cancer-related deaths by 25% by 2025 (MOH, 2016^58^). However, Serenje, being among the pioneer districts in the Central Province, has continued to experience low service uptake. In 2020, only 1,294 out of 4,997 HIV-positive women in the district were screened (SDH, 2021^70^), a relatively low rate compared to other districts in the province. This study aimed to determine factors associated with the underutilization of cervical cancer screening services among HIV-positive women aged 18-69 years in Serenje District.

## METHODS

### Study Design

A cross-sectional study design was utilized. This design allowed the researcher to compare different variables at a specific point in time.

### Study Setting

The study was conducted in Serenje District and involved eight (8) health centers offering Anti-Retroviral Treatment (ART) services. These centers included Muzamani, Mulilima, Nchimishi, Chibale, Kabundi, Serenje HAHC, Kabamba, and Serenje District Hospital.

### Study Population

The study population comprised HIV-positive women aged 18 to 69 years, living with HIV and attending routine HIV treatment services at the selected Public Health Facilities within Serenje District.

#### Inclusion Criteria

Women of 18 to 69 years old, who met the following Criteria; residents of Serenje District, living with HIV positive, present at the time of interview and had lived in Serenje for at least 12 months were included in the study.

#### Exclusion Criteria

The researcher excluded women who were below 18 and above 69 years of age, women who had hysterectomy with removal of the uterine cervix; women with mental or physical limitations and unable to respond to the questions.

### Sample Size determination

The sample size was determined by using the prevalence formula. According to the Zambian fact sheet 2019, a total of 27% women living with HIV utilized the services (UNAIDS, 2019).

**n**= sample size, **Z** (Confidence level of 95%) = 1.96, (Margin of error) **e**= 0.05, prevalence =P. Total sample size was **= 303**

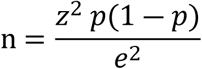

### Sampling Method

Systematic random sampling was used to select respondents at different selected study sites. The calculated sample size (No. 303) was proportionally allocated based on the number of HIV positive women to each facility. Therefore, Probability Proportion to Size (PPS) formula was used as shown below;

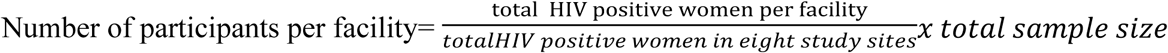

The sampling interval (Kth) for each facility was determined by dividing the average number of HIV positive women attending ART clinic for the past 3 month (sampling frame=N) by the required sample size for each facility (n), therefore the formula;

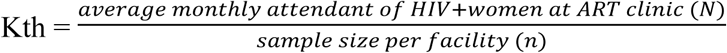

On each day of data collection, the first respondent was randomly selected, and then every Kth eligible respondent was chosen based on their order of arrival until the required sample size for the facility was achieved.

### Data Collection Tool

A researcher-administered questionnaire was used to collect data from 303 respondents aged 18 to 69 years, between 22nd August 2023 and 9th December 2023. The structured questionnaire comprised closed-ended questions to ensure consistency and accuracy in data collection. As the study involved human participants, written informed consent was obtained from all respondents prior to participation. Individuals who declined to participate were not coerced, and all participants were assured protection from emotional harm. The variables collected included age, distance to the nearest health facility, education level, marital status, among others.

### Data Analysis

The data collected from interviews was entered into Microsoft Excel. Data cleaning and error checking were performed before exporting the data to Statistical Software (STATA V.15) for analysis. Descriptive statistics were used to summarize the data for categorical variables, which were reported in frequencies and percentages (e.g., education level, marital status). The Chi-square test was employed to assess the association between the outcome and various categorical independent variables at the unadjusted level, such as the association between the utilization of the screening method and knowledge.

Multiple logistic regression was utilized to predict the relationship between predictors (independent variables) and the predicted variable (dependent variable). Variables with a P-value of less than 0.05 in the bivariate logistic regression analysis (unadjusted level) were included in the multiple logistic regression model. The 95% confidence interval of the odds ratio was computed, and variables with a P-value of less than 0.05 in the multiple logistic regression analysis were considered statistically significant.

## RESULTS

### Characteristics of Study Participants

A total of 303 HIV-positive women participated in the study, of whom 128 (42.24%) utilized cervical cancer screening services, while 175 (57.75%) did not. The majority of participants (84.2%, n=255) were aged between 18 and 40 years, with 105 (41.2%) having utilized screening services, and the remaining 150 (58.8%) not utilizing the services. Most participants (47.5%, n=144) were married, with 63 (43.8%) of them utilizing the services, while 81 (56.3%) did not. Regarding parity, more than half (65.3%, n=198) of the participants had fewer than five children, of whom 80 (40.4%) utilized screening services. Among participants with five or more children (34.7%, n=105), only 48 (45.7%) utilized the services. With respect to average monthly income, the majority (61.4%, n=186) reported earning less than $36.1. Among this group, only 62 (33.3%) utilized the services, while the remaining 124 (66.7%) did not.

Concerning knowledge levels, 217 (71.6%) of the participants had inadequate knowledge about cervical cancer screening, while 86 (28.4%) demonstrated adequate knowledge. Among those with adequate knowledge, 65 (75.6%) utilized the services, compared to only 21 (24.4%) who did not. The majority of respondents (75.6%, n=229) resided more than 5 km from the facilities offering cervical cancer screening services, with only 88 (38.4%) of them utilizing the services. Conversely, among those residing within 5 km of a facility (24.4%, n=74), 40 (54.1%) utilized the services.

Out of the 175 respondents who did not utilize screening services, 151 (86.3%) expressed a negative attitude toward cervical cancer and its screening services, while 24 (13.7%) had a positive attitude but still did not utilize the services. Among the 128 respondents who utilized the screening services, 61 (47.6%) expressed a positive attitude, whereas 67 (52.3%) had a negative attitude despite utilizing the services. Educational attainment also influenced utilization, with 32.3% (n=98) of participants having completed secondary education. Among this group, more than half (52%, n=51) utilized screening services. However, the majority of respondents (50.8%, n=154) had attained only primary education, with 101 (65.6%) not utilizing the services **(Table 1).**

**Table 1:** Demographic Characteristics of respondents.

| Independent variable | Indicator | Utilization of cervical cancer screening |  |  |  |
| --- | --- | --- | --- | --- | --- |
|  |  | Yes | No | Total | Percentage |
| **Age range** | 40+ | 23 | 25 | 48 | (15.8%) |
|  | 30-40 | 51 | 59 | 110 | (36.3%) |
|  | 18-30 | 54 | 91 | 145 | (47.9%) |
|  | Total | 128 | 175 | 303 |  |
| **Marital status** | Married | 63 | 81 | 144 | (47.5%) |
|  | Divorced | 17 | 25 | 42 | (13.9%) |
|  | Widow | 13 | 21 | 34 | (11.2%) |
|  | Single | 35 | 48 | 83 | (27.4%) |
|  | Total | 128 | 175 | 303 |  |
| **Educational level** | Tertiary | 23 | 19 | 42 | (13.9%) |
|  | Secondary | 51 | 47 | 98 | (32.3%) |
|  | Primary | 53 | 101 | 154 | (50.8%) |
|  | No formal education | 1 | 8 | 9 | (3%) |
|  | Total | 128 | 175 | 303 |  |
| **Current employment** | Public/private employed | 25 | 23 | 48 | (15.8%) |
|  | Not employed | 59 | 101 | 160 | (52.8%) |
|  | Self employed | 44 | 51 | 95 | (31.3%) |
|  | Total | 128 | 175 | 303 |  |
| **Parity** | <5/ low parity | 80 | 118 | 198 | (65.3%) |
|  | >5/ grand multiparous | 48 | 57 | 105 | (34.7%) |
|  | Total | 128 | 175 | 303 |  |
| **Income** | >\$108.3/ High | 36 | 12 | 48 | (15.8%) |
| | \$72.2-\$108.3/ Medium | 30 | 39 | 69 | (22.8%) |
| | <\$36.1/ Low | 62 | 124 | 186 | (61.4%) |
|  | Total | 128 | 175 | 303 |  |
| **Knowledge levels on CC screening services** | Adequate Knowledge | 65 | 21 | 86 | (28.4%) |
|  | Inadequate Knowledge | 63 | 154 | 217 | (71.6%) |
|  | Total | 128 | 175 | 303 |  |
| **Attitude of HIV Positive women towards CC screening services** | Positive attitude | 61 | 24 | 85 | (28.1%) |
|  | Negative attitude | 67 | 151 | 218 | (71.9%) |
|  | Total | 128 | 175 | 303 |  |
| **Distance to the cervical cancer screening facility** | Near | 40 | 34 | 74 | (24.4%) |
|  | Far | 88 | 141 | 229 | (75.6%) |
|  | Total | 128 | 175 | 303 |  |
| **Waiting time to access the service** | Less than 30 minutes | 44 | 60 | 104 | (34.3%) |
|  | More than 30 minutes | 84 | 115 | 199 | (65.7%) |
|  | Total | 128 | 175 | 303 |  |

### Utilization of Cervical Cancer Screening Services

This section presents information about the utilization of cervical cancer screening services among HIV-positive women in Serenje District. Out of the 303 participants, 128 (42.24%) reported having utilized the screening services, while the majority (57.76%, n=175) had not. Among the 128 who utilized the services, 76 (59.4%) had been screened within the past two years or more, while 30 (23.4%) underwent screening within the last 12 months **(Fig. 1 and Fig. 2).**

**Figure 1:**
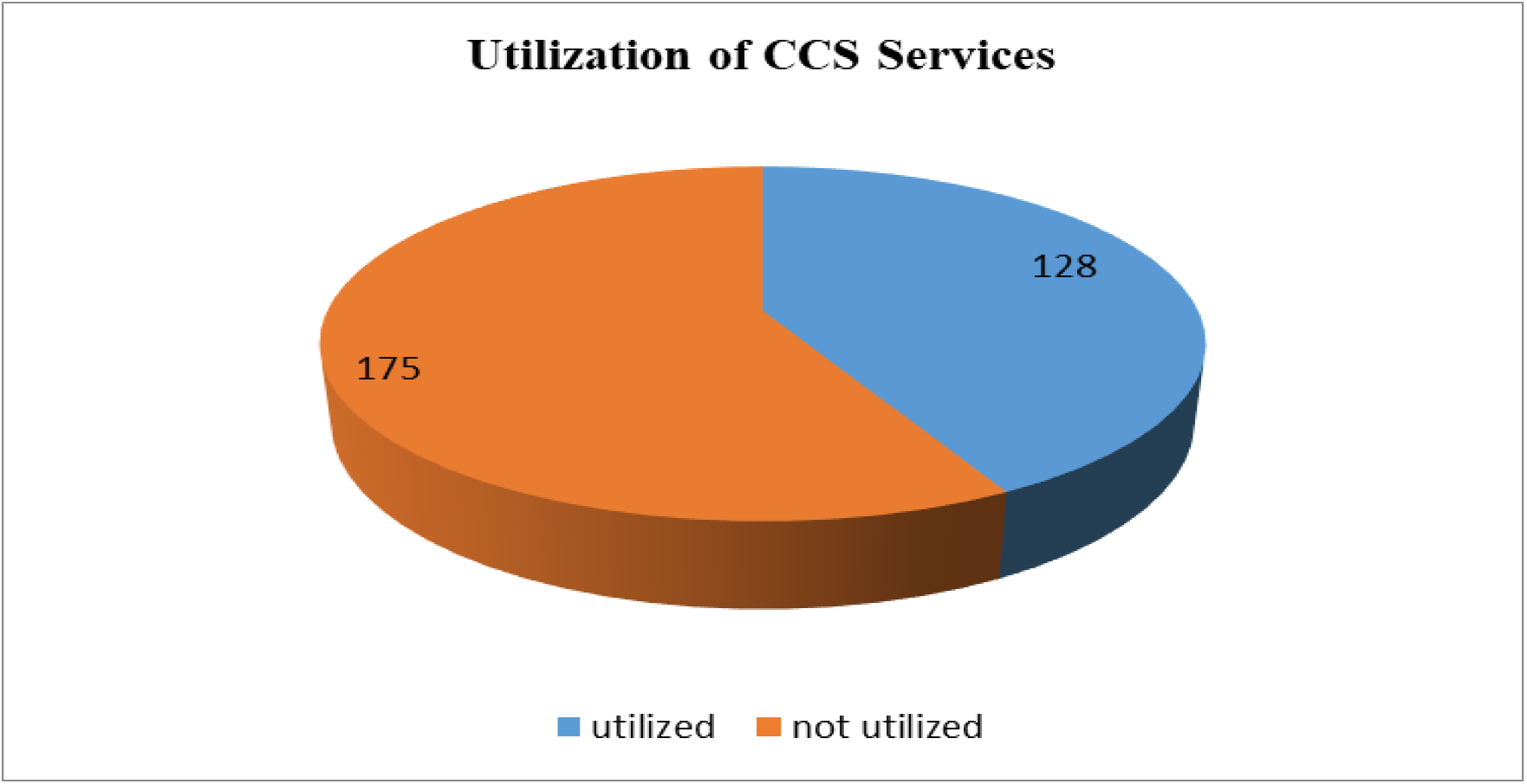
Utilization of Cervical Cancer Screening Services by HIV positive women in Serenje District (n=303).

**Figure 2:**
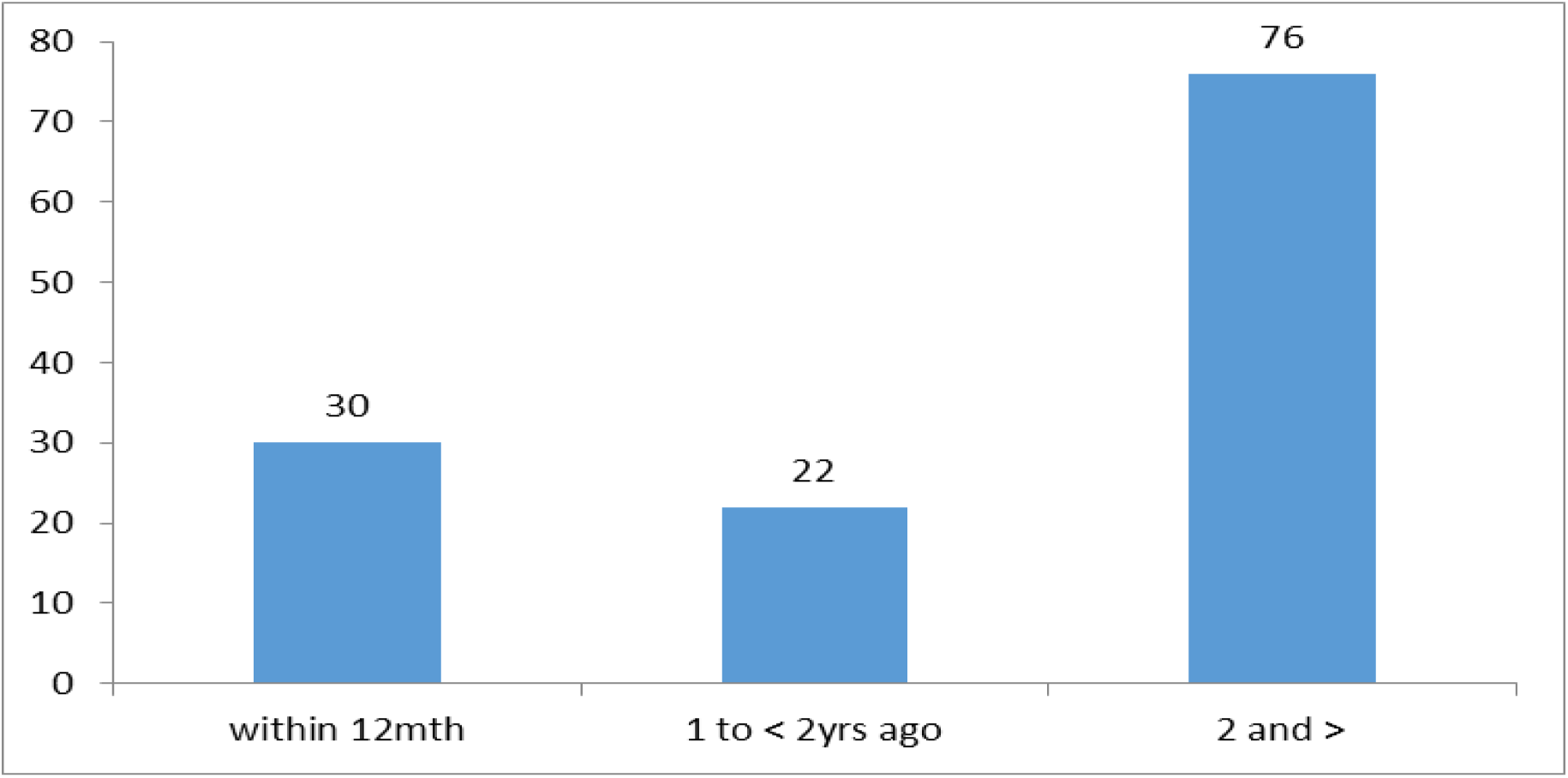
When last the respondent, utilized Cervical Cancer Screening Services

### Facility-Level Characteristics Associated with Underutilization

The study investigated associations between facility-level characteristics and the underutilization of cervical cancer screening services. Variables analyzed included age, marital status, employment status, educational level, parity, income level, knowledge, attitude, cultural beliefs, waiting time, and distance to facilities. To determine whether respondents’ attitudes were positive or negative and their knowledge was inadequate or adequate, a rating scale was used to measure the variables based on their responses.

During bivariate analysis, five variables (educational level, average monthly income, knowledge, attitude, and distance) yielded p-values <0.05 and were included in the multivariable logistic regression analysis. After adjusting for confounders, four variables remained significantly associated with underutilization (p < 0.05) **(Table: 2):**

**Table 2:**
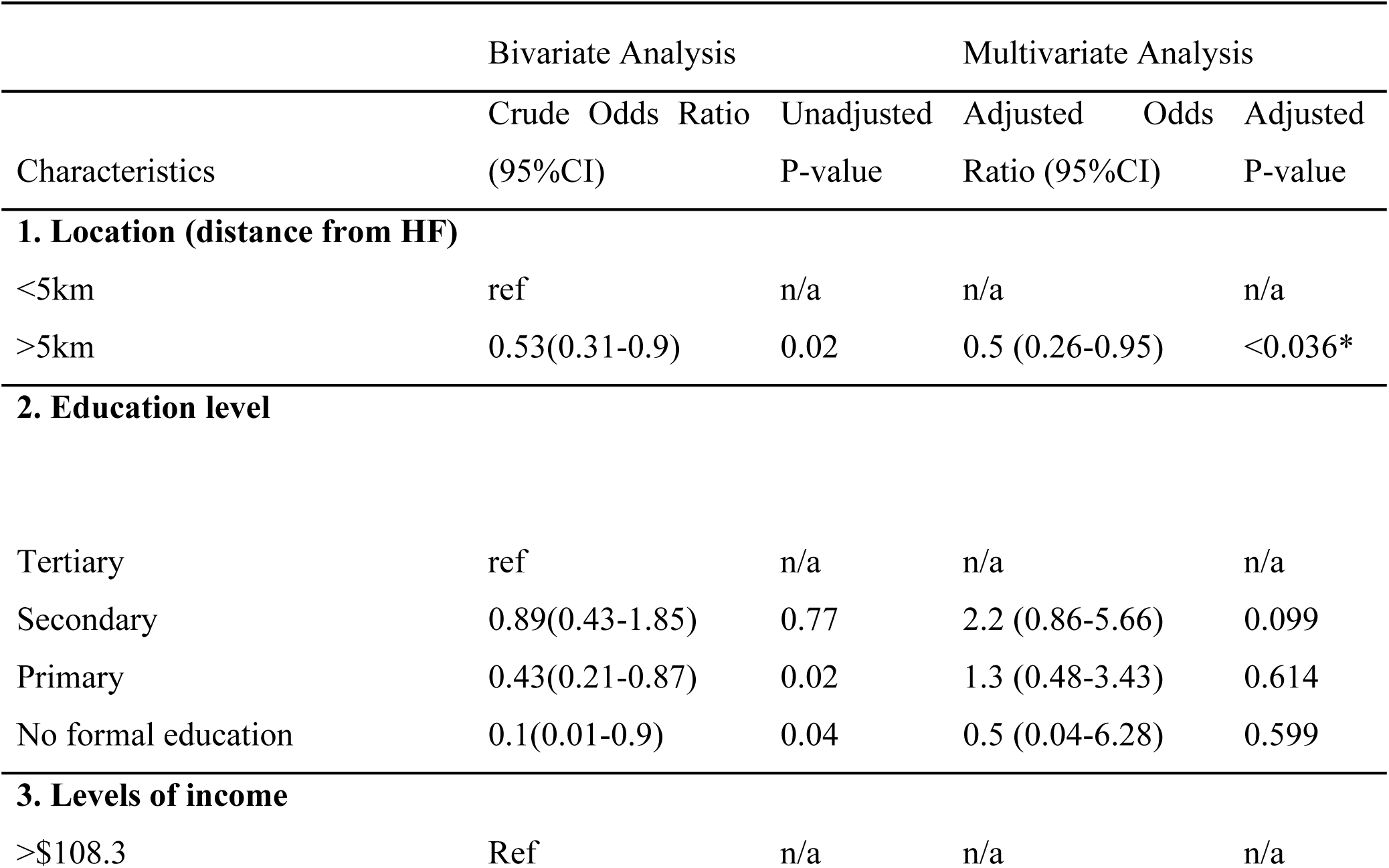
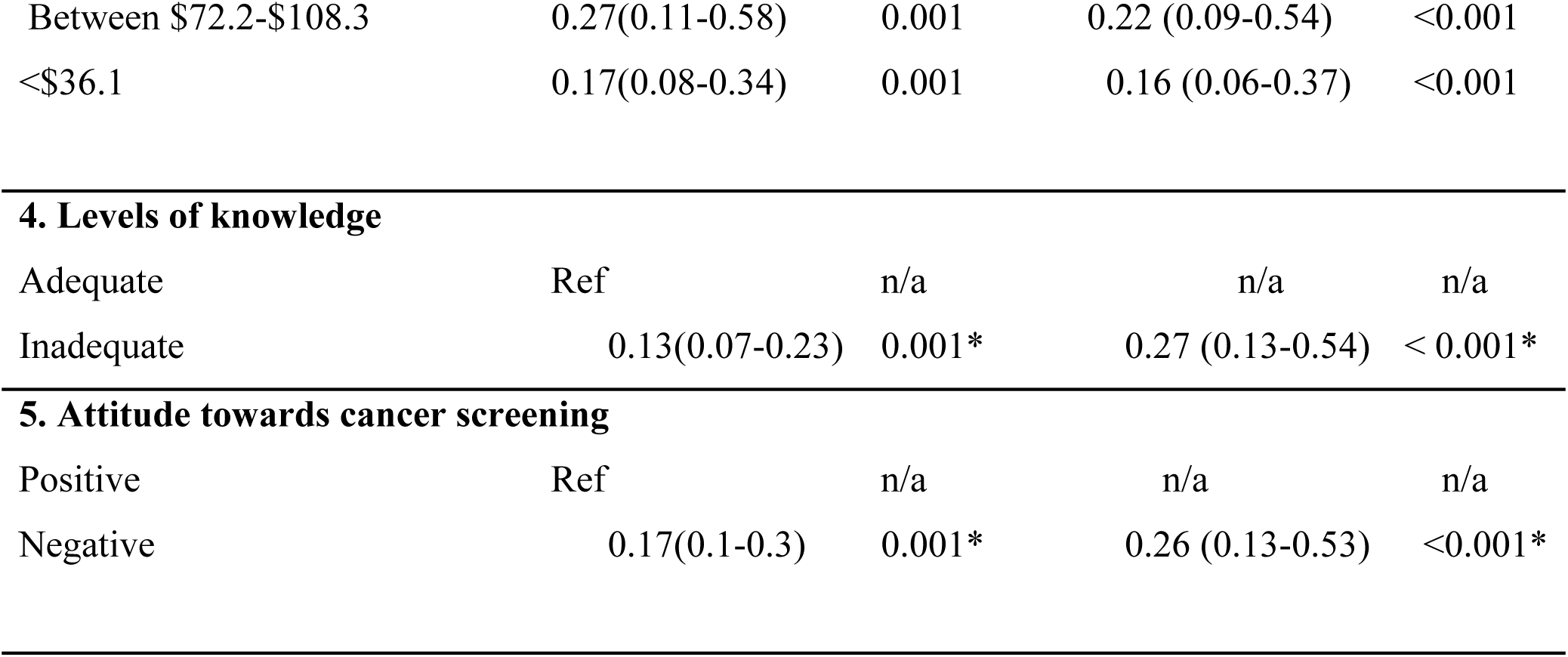
Characteristics associated with underutilization of Cervical Cancer Screening Services.

- **Attitude**: HIV-positive women with a negative attitude were 74% less likely to utilize cervical cancer screening services compared to those with a positive attitude [AOR: 0.26, p<0.001, 95% CI: 0.12–0.53].
- **Knowledge**: Participants with inadequate knowledge were 73% less likely to utilize screening services compared to those with adequate knowledge [AOR: 0.27, p<0.000, 95% CI: 0.13–0.54].
- **Distance**: Women residing more than 5 km from a screening facility were 50% less likely to utilize the services compared to those living within 5 km [AOR: 0.50, p=0.036, 95% CI: 0.25–0.96].
- **Income**: Women with an average monthly income of less than $36.1 were 84% less likely to utilize screening services compared to those earning $108.3 or more [AOR: 0.16, p<0.000, 95% CI: 0.06–0.37]. Similarly, women with an income between $72.2 and $108.3 were 78% less likely to utilize the services compared to those earning $108.3 or more [AOR: 0.22, p<0.001, 95% CI: 0.08–0.54]

## DISCUSSION OF FINDINGS

### Cervical Cancer Screening Services (CCSS)

Study findings revealed an increased utilization rate of cervical cancer screening services in the district among HIV-positive women, with a utilization rate of 42.2%. However, this remains significantly lower than the recommended national target of ensuring 100% annual screening among HIV-positive women. Similar findings have been reported in studies conducted in Ethiopia, where utilization was 41.1% (Assefa et al., 2019^34^), and lower rates were observed in Morocco (Belglaiaa E et al., 2018^02^), Gondar (Nega et al., 2019^35^), and Addis Ababa (Belete, N et al., 2015^36^). The slight increase in utilization within the district may be attributed to the integration of screening services with mobile ART clinics spearheaded by non-governmental organizations operating within the district.

The observed utilization rate, however, falls short compared to studies in countries such as Canada (Leece P et al., 2010^39^), England (Ogunwale A, N et al., 2016^40^), and Thailand (Ploysawang, P., et al., 2021^62^), which reported higher utilization rates. The variation in rates may be attributed to differences in socio-demographic and economic statuses, as well as the health promotion initiatives in these countries. Policies promoting improved access to screening services, nationwide advocacy campaigns, community sensitization, and awareness programs are factors that contribute to higher utilization rates in these regions. Limited distribution of screening facilities in the district may also contribute to the lower utilization rates, unlike countries with universal healthcare systems ensuring widespread availability of primary and specialist care.

### Knowledge of Cervical Cancer Screening Services (CCSS)

Cervical cancer remains a significant health challenge among women in Zambia. This study found that approximately 71.6% of HIV-positive women demonstrated low knowledge about cervical cancer and its screening services. The analysis revealed that HIV-positive women with low knowledge levels were 73% less likely to utilize screening services. Inadequate knowledge in the district may result from the lack of planned information dissemination schedules, insufficient community outreach programs, and limited facility-based sensitization initiatives.

The findings align with other studies reporting that lack of knowledge is a major contributor to low screening service utilization (Adibe et al., 2017^45^; Omowhara et al., 2022^63^; Zulu, 2017^67^; Selmouni et al., 2015^15^; Laranjeira et al., 2013^6^; Ghosh et al., 2020^64^). Factors such as limited time spent between healthcare providers and clients, coupled with the integration of cervical cancer screening into other activities such as ART services, may lead to assumptions that clients are already aware of the program. Increasing awareness campaigns and promoting education at both the community and facility levels could improve knowledge and subsequently utilization rates.

### Attitude of HIV-Positive Women Towards Cervical Cancer Screening Services

The study revealed that 71.9% of respondents exhibited a negative attitude towards cervical cancer screening services, significantly reducing the likelihood of service utilization by 74% [AOR: 0.26 (P<0.001, 95% CI 0.12–0.53)]. A negative attitude may stem from fears, including being attended to by male healthcare providers, concerns about test outcomes, or embarrassment about the procedure. Other contributing factors include misconceptions about the necessity of screening services in the absence of symptoms.

Positive attitudes, on the other hand, are often associated with an understanding of the importance of cervical cancer screening and its benefits, which encourages engagement in preventive measures. Studies have shown that a perceived severity of cervical cancer, along with an understanding of the benefits of screening, motivates women to utilize screening services. Efforts to address these negative perceptions through targeted education and support could improve attitudes and increase utilization.

### Accessibility of Cervical Cancer Screening Services

The results indicated that most HIV-positive women in the study had limited geographical access to screening services, with a significant proportion (75.6%) residing more than five kilometers from the nearest health facility. Similar findings have been reported in other studies where long distances to healthcare facilities and poor transportation infrastructure significantly reduced service utilization (Al-Naggar et al., 2018^65^; Mupepi et al.,2011^18^; Beran et al., 2012^14^). Additionally, it was revealed that HIV-positive women located more than five kilometers away are 50% less likely to utilize cervical cancer screening services [AOR: 0.5 (P<0.036, 95% CI 0.25-0.96)].

In the district, the large catchment area comprising rural communities likely contributed to the low utilization rates. To address this, strategies such as implementing mobile screening camps could bring services closer to communities, reducing the reliance on women traveling long distances to access care. Improved access through community-based services is essential to overcoming the geographical barriers to cervical cancer screening utilization.

### Study Limitations

- The quality of care received during screening was not investigated; therefore, it is unknown whether the care provided was adequate. This would be an important area for future research, as high utilization of cervical cancer screening services is only effective if the care received is of good quality.
- Service-related factors such as staff attitude and the availability of services were not assessed. These factors can significantly influence the utilization of health services.

### Conclusion

The findings from this study indicate inadequate utilization of cervical cancer screening services among HIV-positive women in Serenje District, Zambia. The key determinants associated with the utilization of these services were the distance to healthcare facilities, low levels of knowledge, waiting time, and the attitude of women. These findings should guide policymakers in designing and delivering strategies that target women at high risk.

### Recommendations

- Health personnel at health facilities should intensify targeted and individualized health education on cervical cancer screening services during clinical contacts.
- Community-Based Organizations (CBOs) should be adequately equipped with knowledge about cervical cancer and the importance of screening services, enabling them to sensitize communities effectively.
- The District Health Office should include outreach awareness creation and screening camps to bring services closer to communities, rather than waiting for women to visit health facilities.
- The District Health Office or Provincial Health Office should design and implement information, communication, and education campaigns on cervical cancer screening services through media, women’s groups, and involvement of influential community figures, such as headmen, civic leaders, clergy, and local chiefs.
- The Provincial and District Health Offices should establish mechanisms to enhance monitoring of screening services and the dissemination of information.
- The Government should consider employing more health practitioners to improve staffing levels in selected facilities, reducing client waiting times. This includes hiring midwives, nurses, and clinical officers.
- Increased funding should be allocated to the fight against cervical cancer, enabling health workers to conduct sensitization programs closer to the communities that need them.

## Data Availability

All relevant data are within the manuscript.

## ABBREVIATIONS

CC: Cervical Cancer
CCS: Cervical Cancer Screening
CCSS: Cervical Cancer Screening Services
CDC: Centre of Disease Control and prevention
HPV: Human Papillomavirus
ICC: Invasive Cervical Cancer
MoH: Ministry of Health
VIA: Visual Inspection with Acetate
WHO: World Health Organization
HIV: Human Immunodeficiency Virus
UNAIDS: Joint United Nations programme on HIV/AIDS
LMIC: Low Middle-Income Countries

## Acknowledgement

We acknowledge the University of Zambia, School of Public Health, Department of Community and Family Medicine. Special thanks to the Staff of School of Public Health for the tireless efforts and dedication during the course of Research.

## Ethical and Cultural Consideration

Ethical clearance was obtained from the University of Zambia, Research Ethical Committee (UNZABREC) and authority from the national health research. Further the request was made to the District Health Office in Serenje to allow collecting data from the institution in the district. Since this study involved human subjects, verbal and written consent was sought from the study participants. Those who declined to take part in the research were not coerced and all respondents were free from emotional danger or harm.

## Consent for publication

Not applicable

## Availability of data and material

The datasets used and/or analyzed during the current study are available from the corresponding author and the University of Zambia on reasonable request.

## Authors and affiliations

Department of Community and Family Medicine, School of Public Health. The University of Zambia, Lusaka, Zambia.

Maphious Shindanyi Makina

Phoebe Albina Bwembya

Cosmas Zyambo

Alice Ngoma Hazemba

## Author’s Information

This research was part of a thesis for MSM who was pursuing a Master of Public Health from the University of Zambia, School of Public Health, Department of Community and Family Medicine

## Authors’ contributions

MSM was involved the conceptualization, data curation, data analysis and drafting of the manuscript under the supervision of PAB, CZ and ANH. PAB, CZ and ANH we involved in conceptualization of the study, reviewing and editing the manuscript.

## REFERENCE

1. Stelzle, D., Tanaka, F.L., Lee K.K., Khalil, I.A., Baussano, I., Shah, V.S., McAllister, D. A., Gottlieb, S.L., Klug, S.J., Winkler, A.S., Bray, F., Baggaley, R., Clifford,G.M., Broutet,N. and Shona, D. (2020) ‘Estimates of the global burden of cervical cancer associated with HIV’. Lancet Glob Health, Available at: https://www.thelancet.com/journals/langlo/article/PIIS2214-109X(20)30459-9/fulltext (Accessed: 20. June 2021)

2. Belglaiaa, E., Souho, T., Badaoui, L., Segondy, M., Prétet, J., Guenat, D. and Mougin C. (2018) ‘Awareness of cervical cancer among women attending an HIV treatment centre: a cross-sectional study from Morocco’. BMJ Publishing Group. Available at: https://bmjopen.bmj.com/content/8/8/e020343 (Accessed: 16. May 2020)

3. Mwanahamuntu, H.M., Sahasrabuddhe, V., Blevins, M., Kapambwe, S., Shepherd, B.E., Chibwesha, C., Pfaendler, K.S., Mkumba, G., Vwalika, B., Hicks, M.L., Stringer, J. S. and Groesbeck, P.P. (2013) ‘Utilization of Cervical Cancer Screening Services and Trends in Screening Positivity Rates in a ‘Screen-And-Treat’ Program Integrated with HIV/AIDS Care in Zambia’. PLOS ONE, Available at: https://www.ncbi.nlm.nih.gov/pmc/articles/PMC3776830/#!po=1.78571 (Accessed: 04 April 2021)

4. Fort, V.K., Siegler, A.J., Ault, K. and Rochat, R. (2011) ‘Barriers to cervical cancer screening in Mulanje, Malawi: a qualitative study’. Pub Med. Available at: https://www.ncbi.nlm.nih.gov/pmc/articles/PMC3063659/ (Accessed: 20 April 2021)

5. Di1, J., Rutherford, S., Wu, J., Song, B., Ma, L., Chen, J. and Chu, J. (2016) ‘Knowledge of Cervical Cancer Screening among Health Care Workers providing services across different Socio-economic Regions of China’, National library of medicine, Pub Med. Available at: https://pubmed.ncbi.nlm.nih.gov/27356719/ (Accessed: 20 May 2021)

6. Laranjeira, A. C. (2013) ‘Portuguese women’s knowledge and health beliefs about cervical cancer and its screening’. National library of medicine. Pub Med. Available at: 03 April 2020. https://pubmed.ncbi.nlm.nih.gov/23461350/ (Accessed: 03 April 2021)

7. Matangaidze, O. (2015) ‘Knowledge, attitude and practices of HIV infected women on cervical cancer screening at Musiso mission hospital, Masvingo province, Zimbabwe’. University of Limpopo, Available at: http://ulspace.ul.ac.za/ (Accessed: 05 April 2021)

8. Twinomujuni, C., Nuwaha, F. and Babirye, J. N. (2015) ‘Understanding the Low level of cervical cancer screening in Masaka Uganda using the ASE model: a community-based survey’. National library for medicine. Available at: https://pubmed.ncbi.nlm.nih.gov/26030869/ (Accessed: 02 April 2021)

9. Mutambara, J., Mutandwa, P., Mahapa, M., Chirasha, V., Nkiwane, S. and Shangahaidon, T. (2017) ‘Knowledge, attitudes and practices of cervical cancer screening among women who attend traditional churches in Zimbabwe’. Journal of Cancer Research and Practice. Available at: https://www.sciencedirect.com/science/article/pii/S2311300616301641 (Accessed: 05 May 2021)

10. Bulto, G.A, Demmissie, D. B. and Daka K. B. (2016) ‘Knowledge about Cervical Cancer, Screening Practices and associated factors among Women Living with HIV in Public Hospitals of West Shoa Zone, Central Ethiopia’. Journal of Women’s Health Care (Accessed: 5 May 2021)

11. Ngugi, W.C., Boga, H., Muigai, A.W.T., Wanzala, P. and Mbith, J.N. (2012) ‘Factors affecting uptake of cervical cancer early detection measures among women in Thika, Kenya,’ National library for medicine, Pub Med. Available at: https://pubmed.ncbi.nlm.nih.gov/22681745/ (Accessed: 05 April 2021)

12. Liu, T., Shunping, L, Ratcliffe, J. and Chen, G. (2017) ‘Assessing Knowledge and Attitudes towards Cervical Cancer Screening among Rural Women in Eastern China’. International journal of Environmental research and public health, p-1

13. Vhuromu, E.N, Daniel T. G, Maputle, M.S, Lebese, R.T and Okafor, B. (2018) ‘Utilization of Cervical Cancer Screening Services among Women in Vhembe District, South Africa: A Cross-Sectional Study’, The Open Public Health Journal. (Accessed: 10 May 2020)

14. Lyimo, F. S and Beran, T. N. (2012) ‘Demographic, knowledge, attitudinal, and accessibility factors associated with uptake of cervical cancer screening among women in a rural district of Tanzania: three public policy implications’. BMC Public Health

15. Selmouni, F., Zidouh, A and Alvarez-Plaza, C (2015) Perception and satisfaction of cervical cancer screening by Visual Inspection with Acetic acid (VIA) at Meknes-Tafilalet Region, Morocco: a population-based cross-sectional study. BMC Women’s Health 15, 106 (2015). 10.1186/s12905-015-0268-03.

16. Banda, L., Nyirongo, T. and Muntanga, M. (2019) ‘Cancer – An Emerging Health Problem: The Zambian Perspective’. Health Press Zambia Bull. p. 3

17. Gan, D. E. H. and Dahlui, M. (2013) ‘cervical screening uptake and its predictors among rural women in Malaysia’. Pub med.

18. Mupepi, C. S., Sampselle, C.M. and Johnson, T.R.B. (2011) ‘Knowledge, Attitudes, and Demographic Factors Influencing Cervical Cancer Screening Behaviour of Zimbabwean Women’. Journal of women’s health (Accessed: 02 June 2020)

19. Venturas, C and Umeh, K. (2017) ‘Health professional feedback on HPV vaccination roll-out in a developing country’. Pub Med

20. Kalubula, M., Shen, H., Makasa M & Liu, L. (2018) ‘Epidemiology of cancers in Zambia; A significant variation in cancer incidence and prevalence across the nation’. Pub Med. (Accessed: 20 May 2021)

21. Bray F., Ferlay J., Soerjomataram I., Siegel R.L., Torre L.A. and Jemal A. (2018) ‘Global Cancer Statistics 2018: GLOBOCAN estimates of incidence and mortality worldwide for 36 cancers in 185 countries. A Cancer Journal for Clinicians. Available at: http://gco.iarc.fr/ (Accessed: 20 April 2021)

22. Twinomujuni, C., Nuwaha, F. and Babirye J.N. (2015) ‘Understanding the Low level of cervical cancer screening in Masaka Uganda using the ASE model: a community-based survey’. National library of medicine. Available at: https://pubmed.ncbi.nlm.nih.gov/26030869/ (Accessed: 20 May 2021)

23. Ramathuba, D.U, Ngambi, D, Khoza, L.B & Ramakuela, N.J (2016), ‘Knowledge, attitudes and practices of women regarding cervical cancer prevention at Thulamela Municipality of Vhembe District in Limpopo Province’. Health Edu Recrea Dance

24. Pandey, A. R. and Karmacharya. E. (2017) cervical cancer screening behavior and associated factors among women of Ugrachandi Nala, Kavre, Nepal, national library for medicine, Pub Med. Available at: https://pubmed.ncbi.nlm.nih.gov/28927464/ (Accessed: 20 April 2021)

25. UNAIDS (2020) ‘THEMATIC SEGMENT: Cervical cancer and HIV–– addressing linkages and common inequalities to save women’s lives’. Available at: https://www.unaids.org/sites/default/files/media_asset/PCB47_Thematic_Segment_BNEN.pdf (Accessed: 05 May 2021)

26. Mtengezo, J. (2019) ‘Knowledge and Attitudes Regarding Cervical Cancer Screening Knowledge and Attitudes Regarding Cervical Cancer Screening Among Women Living with HIV/AIDs in Malawi: A Cross-sectional Among Women Living with HIV/AIDs in Malawi: A Cross-sectional Study’, Boston University of Massachusetts. (Accessed: 20 May 2021)

27. World Health Organization (2020) WHO Global strategy to accelerate the elimination of cervical cancer a s a public health problem. Available at: https://www.uicc.org/resources/whos-global-strategy-accelerate-elimination-cervical-cancer-public-health-problem. (Accessed: 20 May 2021)

28. Project concern international (2020) ‘gaining ground on cervical cancer in Zambia; A global communities partner’. Available at: https://www.pciglobal.org/gaining-ground-on-cervical-cancer-in-zambia/ (Accessed: 06 June 2021)

29. Zambia Fact Sheet (2018) ‘Joint United Nations Programme on HIV/AIDS’. Available at: https://www.unaids.org/sites/default/files/country/documents/ZMB_2020_countryreport.pdf (Accessed: o3 June 2021)

30. World Health Organisation (2020) ‘WHO Costing the National Strategic Plan on Prevention and Control of Cervical Cancer: Zambia, 2019—2023’. Available at: 3/4/2021, https://www.who.int/publications/m/item/costing-the-national-strategic-plan-on-prevention-and-control-of-cervical-cancer-zambia-2019-2023 (Accessed: 20 May 2021)

31. McFarland, D.M., Gueldner, S.M and Mogobe, K.D (2016) Integrated Review of Barriers to Cervical Cancer Screening in Sub-Saharan Africa. J Nurs Scholarsh. 2016 Sep;48(5):490–8. doi: 10.1111/jnu.12232. Epub 2016 Jul 19. PMID: 27434871. (Accessed: 20 May 2021)

32. Morema, E. N., Atieli, H.E., Onyango, R.O., Omondi, J. H. and Ouma,C. (2014) ‘Determinants of Cervical screening services uptake among 18–49-year-old women seeking services at the Jaramogi Oginga Odinga Teaching and Referral Hospital, Kisumu, Kenya’. BMC Health Services Research, p. 1

33. Mwaka, A. D., Orach, C. G., Were, E.M., Lyratzopoulos, G., Wabinga, H. and Roland, M. (2015) ‘Awareness of cervical cancer risk factors and symptoms: cross-sectional community survey in post-conflict northern Uganda’. Health Expect (Accessed: 20 April 2021)

34. Assefa, A.A. (2019) Cervical cancer screening service utilization and associated factors among HIV positive women attending adult ART clinic in public health facilities, Hawassa town, Ethiopia: a cross-sectional study.

35. Nega, A.D., Woldetsadik., M.A. and Gelagay, A.A. (2019) Low uptake of cervical cancer screening among HIV positive women in Gondar University referral hospital. BMC Health Services Research 19:847 Page 10 of 11 Northwest Ethiopia: cross-sectional study design. BMC Womens Health. 2018;18(1):1–718

36. Belete, N., Tsige, Y. and Mellie, H. (2015) ‘Willingness and acceptability of cervical cancer screening among women living with HIV / AIDS in Addis Ababa, Ethiopia: a cross sectional study’. Gynecol Oncol Res Pract. 2015:4–9. Available from. 10.1186/s40661-015-0012-3

37. Wanyenze, R.K., Baptist, J., Beyeza-kashesya, J., Mugerwa, S. and Arinaitwe, J. (2017) ‘Uptake and correlates of cervical cancer screening among HIV-infected women attending HIV care in Uganda’. Global Health Action, available from 10.1080/16549716.2017.1380361

38. Erku, D.A., Netere, A.K., Mersha, A.G. and Abebe, S.A. (2017) ‘Comprehensive knowledge and uptake of cervical cancer screening is low among women living with HIV / AIDS in Northwest Ethiopia’. Gynecol Oncol Res Pract. 2017; 4(20):1–7]

39. Leece, P., Kendall, C., Touchie, C., Angel, J.B., Jaffey, J. and Pottie K. (2010) Cervical cancer screening among HIV-positive women Recherche les femmes VIH positives. Can Fam Physician. 2010; 56:425–31

40. Ogunwale, A.N., Coleman, M.A., Sangi-Haghpeykar, H., Valverde, I., Montealegre, J. and Jibaja-Weiss, M.A.M. (2016) ‘Assessment of factors impacting cervical cancer screening among low-income women living with HIV-AIDS’. AIDS Care. 2016; 28(4):491–4.

41. Stuardo, V., Agustí, C., Casabona, J. and Study, B.H. (2013) ‘Low prevalence of cervical Cancer screening among HIV-positive women in Catalonia (Spain) AIDS & Clinical Research’. J AIDS Clin Res. 2013; S3 (004):3–5]

42. Rosser, J.I., Njoroge, B. and Huchko, M.J. (2015) ‘Cervical Cancer Screening Knowledge and Behavior among Women Attending an Urban HIV Clinic in Western Kenya’. J Cancer Educ. 2015; 30 (3):567

43. Oba, S., Toyoshima, M. and Ogata, H. (2017) ‘Association of Cervical Cancer Screening with Knowledge of Risk Factors, Access to Health Related Information, Health Profiles, and Health Competence Beliefs among Community Dwelling Women in Japan’. Asian Pac J Cancer Prev. 2017; 18(8):2–3] and China

44. Leung, S.S.K. and Leung, I. (2010) ‘Cervical Cancer Screening: knowledge, health perception and attendance rate among Hong Kong Chinese women’. Int J Women’s Health. 2010; 2:221–8].

45. Adibe, M.O. and Aluh, D.O. (2017) ‘Awareness, knowledge and attitudes towards cervical cancer amongst hiv-positive women receiving care in a Tertiary Hospital in Nigeria’. J Cancer Educ 2017. doi: 10.1007/s13187-017-1229-0. Epub ahead of print 5 May 2017.

46. Maree, J.E. and Moitse, K.A. (2015) ‘Exploration of knowledge of cervical cancer and cervical cancer screening amongst HIV-positive women’. Curationis 2014; 37:1209. 39.

47. Idowu, A., Olowookere, S.A., Fagbemi, A.T. and Ogunlaja, O.A. (2016) ‘Determinants of cervical cancer screening uptake among women in Ilorin, North Central Nigeria: a community-based study’. Journal of cancer epidemiology.

48. Bayu, H., Berhe, Y., Mulat, A and Alemu, A. (2015) ‘Cervical Cancer Screening Service Uptake and Associated Factors among Age Eligible Women in Mekelle Zone, Northern *Ethiopia*’. A Community Based Study Using Health Belief Model. PLOS/ONE. 2016; 11 (3):1–13.

49. Geremew, A.B., Gelagay, A.A. and Azale, T. (2018) ‘Uptake of pre cervical cancer screening service and associated factors among women aged 30-49 years in Finote Selam town Northwest Ethiopia’. Int J Collab Res Intern Med Public Heal. 2018; 10 (2):829–42

50. Cervical cancer screening in Canada: Environmental scan (2018) ‘Canadian Partnership against cancer’. Available at: https://www.partnershipagainstcancer.ca/topics/cervical-cancer-screening-environmental-scan-2018/ (Accessed: 04 April 2021)

51. Ferlay, J., Ervik, M. and Lam, F. (2018) ‘Global Cancer Observatory: Cancer Today. Lyon, France: International Agency for Research on Cancer’. (Accessed: 20 May 2021)

52. Anderson, R.M (1995) ‘Revisiting the behavioural model and access to medical care; does it matter?’ (Accessed: 20 May 2021)

53. Gossa, W. and Fetters, M.D. (2020) ‘How should cervical cancer prevention be improved in LMICs’, AMA Journal of Ethics. (Accessed: 07 April 2021)

54. Adeyodi, A. O. and Lim, J.N.W. (2017) ‘Barriers to utilization of cervical cancer screening in Sub Sahara Africa: a systematic review’. Pub Med. (Accessed: 23 May 2021)

55. World Health Organisation (2020) Human papilloma virus and cervical cancer report. Available at: https://www.who.int/news-room/fact-sheets/detail/human-papillomavirus-(hpv)-and-cervical-cancer (Accessed: 05 May 2021)

56. World Health Organisation (2020) ‘WHO launch of the global strategy to accelerate the elimination of cervical cancers’. Available at: https://www.who.int/news/item/17-11-2020-a-cervical-cancer-free-future-first-ever-global-commitment-to-eliminate-a-cancer#:∼:text=WHO’s%20Global%20Strategy%20to,million%20related%20deaths%20by%202050. (Accessed: 20 April 2021)

57. World Health Organisation (2020) ‘Accelerate Cervical Cancer Elimination Initiative’. Available at: https://www.thelancet.com/journals/lanonc/article/PIIS1470-2045(20)30729-ed0important%20steps%3A (Accessed: 04 May 2021)

58. MOH Zambia (2016) ‘National Cancer Control Strategic Plan 2016-2021’.

59. UNAIDS (2019) ‘the little-known links between cervical cancer and HIV’. (Accessed: 20 May 2021)

60. World Health Organisation (2020) ‘Cancer Today: data visualization tools for exploring the global cancer burden in 2020’. Available at: https://gco.iarc.fr/today/home (Accessed: 04 May 2021)

61. Maddux JE, Rogers RW. Protection motivation and self-efficacy: A revised theory of fear appeals and attitude change. Journal of experimental social psychology. 1983;19 (5):469–794]

62. Ploysawang, P., Rojanamatin, J., Prapakorn, S., Jamsri, P., Pangmuang, P., Seeda, K and Sangrajrang, S (2021) ‘National Cervical Cancer Screening in Thailand. Asian’ Pac J Cancer Prev. 2021 Jan 1;22(1):25–30. doi: 10.31557/APJCP.2021.22.1.25. PMID: 33507675; PMCID: PMC8184188

63. Omowhara, O.B., Soter, S.A and Banjo, A. A (2022) ‘Cervical cancer screening awareness and uptake among under screened women in rural Nigerian community’. The Nigerian health journal; Volume 22

64. Ghosh, S., Mallya, S.D., Shetty, R, S., Pattanshetty, S.M., Pandey, D., Kabekkodu, S.P., Satyamoorthy, K and Kamath, V.G (2020) ‘Knowledge, Attitude and Practices Towards Cervical Cancer and its Screening Among Women from Tribal Population: A Community-Based Study from Southern India’. J Racial Ethn Health Disparities. 2021 Feb;8(1):88–93. doi: 10.1007/s40615-020-00760-4. Epub 2020 Apr 24. PMID: 32333376; PMCID: PMC7853713.

65. Al-Naggar, R.A., Low, W.Y and Isa, Z.M (2010) ‘Knowledge and barriers towards cervical cancer screening among young women in Malaysia’. Asian Pac J Cancer Prev. 2010;11(4):867–73. PMID: 2113359

66. Devarapalli, P., Labani, S., Nagarjuna, N., Panchal, P and Asthana S (2018) Barriers affecting uptake of cervical cancer screening in low and middle income countries: A systematic review. Indian J Cancer. 2018 Oct-Dec;55(4):318–326. doi: 10.4103/ijc.IJC_253_18. PMID: 30829264.

67. Zulu, W (2017) ‘Factors influencing utilization of cervical cancer screening services by women at selected clinics of Lusaka urban District of Zambia’. The University of Zambia (Accessed: 04 May 2021)

68. Ministry of Health (2020) ‘Cervical Cancer Screening Guidelines’. Zambia (Accessed: 14 May 2020)

69. Sung, H., Ferlay, J., Siegel, R.L., Laversanne, M., Soerjomataram, I., Jemal A and Bray F (2021) ‘Global cancer statistics 2020: GLOBOCAN estimates of incidence and mortality worldwide for 36 cancers in 185 countries’, CA A Cancer. J. Clin. J. Clin. 2021 doi: 10.3322/caac.21660.

70. Serenje District Health Office (2021) Health Management Information System

